# LigaNET: A multi-modal deep learning approach to predict the risk of subsequent anterior cruciate ligament injury after surgery

**DOI:** 10.1101/2023.07.25.23293102

**Authors:** Mo Han, Mallika Singh, Davood Karimi, Jin Young Kim, Sean W. Flannery, BEAR Trial Team, Kirsten Ecklund, Martha M. Murray, Braden C. Fleming, Ali Gholipour, Ata M. Kiapour

**Affiliations:** Department of Orthopaedic Surgery, Boston Children’s Hospital, Harvard Medical School, 300 Longwood Avenue, Boston, MA 02115, USA; Department of Radiology, Boston Children’s Hospital, Harvard Medical School, 300 Longwood Avenue, Boston, MA 02115, USA; Department of Orthopaedics, Warren Alpert Medical School of Brown University, Rhode Island Hospital, 1 Hoppin St, Providence RI 02903, USA

## Abstract

Anterior cruciate ligament (ACL) injuries are a common cause of soft tissue injuries in young active individuals, leading to a significant risk of premature joint degeneration. Postoperative management of such injuries, in particular returning patients to athletic activities, is a challenge with immediate and long-term implications including the risk of subsequent injury. In this study, we present LigaNET, a multi-modal deep learning pipeline that predicts the risk of subsequent ACL injury following surgical treatment. Postoperative MRIs (n=1,762) obtained longitudinally between 3 to 24 months after ACL surgery from a cohort of 159 patients along with 11 non-imaging outcomes were used to train and test: 1) a 3D CNN to predict subsequent ACL injury from segmented ACLs, 2) a 3D CNN to predict injury from the whole MRI, 3) a logistic regression classifier predict injury from non-imaging data, and 4) a multi-modal pipeline by fusing the predictions of each classifier. The CNN using the segmented ACL achieved an accuracy of 77.6% and AUROC of 0.84, which was significantly better than the CNN using the whole knee MRI (accuracy: 66.6%, AUROC: 0.70; P<.001) and the non-imaging classifier (accuracy: 70.1%, AUROC: 0.75; P=.039). The fusion of all three classifiers resulted in highest classification performance (accuracy: 80.6%, AUROC: 0.89), which was significantly better than each individual classifier (P<.001). The developed multi-modal approach had similar performance in predicting the risk of subsequent ACL injury from any of the imaging sequences (P>.10). Our results demonstrate that a deep learning approach can achieve high performance in identifying patients at high risk of subsequent ACL injury after surgery and may be used in clinical decision making to improve postoperative management (e.g., safe return to sports) of ACL injured patients.

## Introduction

Traumatic injuries of joint connective tissues (e.g., ligament tears) are among the most common musculoskeletal conditions in adolescents and adults participating in physically demanding activities such as sports and military operations (1). A ruptured anterior cruciate ligament (ACL) is one of the most common and devastating connective tissue injuries, primarily affecting young, active individuals such as athletes and soldiers (2-4). The current standard of care for the treatment of an ACL injury is ACL reconstruction in which the injured ligament is replaced by a graft of tendon harvested from the patient or a donor. ACL reconstruction has shown promising results in restoring the gross stability of a symptomatic ACL-deficient knee; however, it is also associated with high rate of secondary injuries, particularly in the adolescent population (5, 6). A young patient who returns to sport within 1 year is 15 times more likely to suffer a second ACL injury than a healthy control with no history of a knee injury (7). This injury risk remains elevated in the first two years of returning to activity, when a patient with ACL injury is approximately 6 times more likely to sustain a second injury than an uninjured counterpart (8). These reinjuries can lead to accelerated knee arthrosis, and ultimately, reduced activity and early disability (9-11). All of these may in turn impose a significant burden to society, financially as well as socially (12, 13). These statistics highlight the need to optimize postoperative care and return-to-sports guidance.

Development of an effective patient-specific post-operative care plan and establishing when it is safe to return to sport following an ACL surgery is one of the most challenging decisions that may be made by a sports medicine team. There has been a recent growth in research identifying return-to-sports criteria with the aim to reduce the risk of a second ACL injuries (14, 15). The current tools for clinical decision making include a battery of clinical examination tests (e.g., range of motion, muscle strength), knee functional assessments (e.g., hop testing, balance testing), and patient reported outcomes (e.g., questionnaires). Despite promises in lowering the risk of graft injuries (14, 15), and the ability to assess overall knee function and neuromuscular performance, the current tools fail to directly evaluate the healing ACL graft properties (16, 17). Moreover, these measures are often influenced by factors unrelated to the ACL structure. For example, physical examinations of the knee (e.g., the Lachman and pivot shift tests) can be influenced by the injury or hypertrophy of secondary stabilizers of the knee (18, 19), as well as age (20), sex (20), and bony anatomy (21), and are prone to observer bias. Functional testing can be influenced by the quality of the rehabilitation program, patient compliance, and/or fear of reinjury (22, 23). Likewise, patient-reported outcomes after ACL surgery have been shown to be influenced by self-esteem levels (24), body mass index (25), and smoking (25). The relatively low sensitivity and specificity of the current approaches to track the status of healing ACL also pose serious challenges in assessment of the efficacy of the new surgical techniques and to guide their post-operative care, considering the significant lack of clinical data on these recently developed techniques (15).

Alternatively, magnetic resonance imaging (MRI) offers a non-invasive approach to directly assess the structural integrity of the healing ACL following surgery (16, 17). MRI has been previously used in preclinical (26-34) and clinical (35-48) studies evaluating postoperative changes in healing ACL after surgery. MRI-based signal intensity and T_2_* relaxation times are among the most used quantitative MRI parameters to evaluate healing ACL properties (16, 17). Our group, among others, have shown signal intensity is associated with graft mechanical and histological properties after ACL reconstruction, in rabbit, porcine and sheep knees (26-34, 49-51). We have also shown that T_2_* relaxation time is associated with the tensile structural properties of the surgically treated porcine knees (27-30, 33). Following ACL surgery, both signal intensity and T_2_* relaxation time have been shown to decrease over time, indicating that the healing ACL is becoming more organized and stronger with time (27, 39, 43, 52). These previous studies have used regional or overall signal intensity values or T_2_* relaxation times to predict the ACL structural properties and outcomes of surgery. However, the methods used do not capture the complex 3-dimentional distribution of the ACL signal intensity or T_2_* relaxation time, which change following surgery. Computational deep learning approaches are well suited for this type of application as they are capable of identifying layers of features and complex patterns (53, 54). Such tools have recently been used in several musculoskeletal studies including cartilage lesion detection (55, 56), knee osteoarthritis prediction (57, 58), knee injury classification (59), and ligament automatic segmentation from MRI (60, 61).

In this work, we present LigaNET, a new multi-modal deep learning algorithm to predict the risk of subsequent ACL injury following surgical treatment based on a combination of imaging and non-imaging predictors. We hypothesized that a multi-modal deep learning algorithm that uses both imaging features and non-imaging data, would more accurately predict the risk of subsequent ACL surgery than its counterparts based on either imaging features or non-imaging data. LigaNET differs conceptually from previous MRI-based techniques as it uses detailed features of knee MRI (e.g., signal intensity distribution) at multiple levels, to predict the risk of subsequent injury, instead of solely focusing on average ACL signal intensity or T2* relaxation values. A Convolutional Neural Network (CNN) within LigaNET was first trained and validated to extract structural features of intact ACLs, surgically treated ACLs, and ACL grafts from MRI. We then used this feature extractor to develop and validate two additional 3D CNN classifiers to predict risk of subsequent ACL injury using either isolated segmented ACLs and ACL grafts from MR or whole-knee MR images. Finally, we used various combinations of imaging (i.e., isolated MRI-segmented ACL and whole-knee MRI) and non-imaging clinical predictors to train and validate LigaNET as an integrated multimodal classification algorithm to predict the subsequent ACL injury risk.

We systematically assessed the performance of LigaNET and alternative models using data from a series of FDA-approved clinical trials of ACL surgical treatments (ACLR: ACL reconstruction, and BEAR: Bridge-Enhanced ACL Restoration). Model performance was assessed and compared based on overall accuracy, sensitivity, and specificity, along with areas under the receiver operating characteristic (AUROC) and precision-recall curves (AUPRC). We also used occlusion maps to better understand the features that the model used to assign the classifications. The occlusion maps as well as standard performance metrics (i.e., accuracy, AUROC and AUPRC) were used to assess if the MRI sequence had a major effect on models’ ability to predict the risk of ACL injury. All endpoints were selected prior to data collection. The study was approved by the Boston Children’s Hospital Institutional Review Board, and all methods were carried out in accordance with the approved study protocol.

## Materials and Methods

### Study Design

The goal of this study was to develop and conduct a proof-of-concept evaluation of a multi-modal deep learning pipeline to predict the risk of subsequent ACL injury based on a combination of imaging and non-imaging predictors. We first trained and validated a CNN feature extractor to study structural features of intact ACLs, surgically treated ACLs and ACL grafts from MRI. We then used this feature extractor to develop and validate two additional 3D CNN classifiers to predict risk of subsequent ACL injury using either isolated segmented ACLs and ACL grafts from MR or whole-knee MR images. Ultimately, we tested various combinations of imaging (i.e., isolated MRI-segmented ACL and whole-knee MRI) and non-imaging clinical predictors to train and validate a multi-modal pipeline that can predict the subsequent ACL injury risk. Model performance was assessed and compared based on overall accuracy, sensitivity, and specificity, along with areas under the receiver operating characteristic (AUROC) and precision-recall curves (AUPRC). We finally used occlusion maps to better understand the features that the model used to assign the classifications. The occlusion maps as well as standard performance metrics (i.e., accuracy, AUROC and AUPRC) were used to assess if the MRI sequence had a major effect on models’ ability to predict the risk of ACL injury. All endpoints were selected prior to data collection. The study was approved by the Boston Children’s Hospital Institutional Review Board, and all methods were carried out in accordance with the approved study protocol.

### Participants and Inclusion/Exclusion Criteria

The comprehensive data sets from three IRB and FDA approved clinical trials of ACL surgery (BEAR I: n=20, NCT02292004 (62); BEAR II: n=100, NCT02664545 (63); BEAR III: n=39; NCT03348995(45)) were used. The cohort included 69 males and 90 females with an average age of 19.8 ± 5.2 years (range: 14 – 36). All patients granted their written informed consent prior to participating. All patients presented with a complete ACL tear, were less than 45 days from injury, had closed physes, and had at least 50% of the length of the ACL attached to the tibia (as determined from a pre-operative MR image). From 159 patients, 114 were treated bridge-enhanced ACL repair (BEAR) and 45 were treated with ACL reconstruction (ACLR) using hamstrings (n=43) or bone-patellar tendon-bone (n=2) autografts. Patients were excluded from enrollment if they had a history of prior ipsilateral knee surgery, history of prior knee infection, or had risk factors that could adversely affect ligament healing (nicotine/tobacco use, corticosteroids in the past six months, chemotherapy, diabetes, inflammatory arthritis). Patients were also excluded if they had a displaced bucket handle tear of the medial meniscus requiring repair. All other meniscal injuries were included. Patients were also excluded if they had a full thickness chondral injury, a grade III MCL injury, a concurrent complete patellar dislocation, or an operative posterolateral corner injury. Detailed descriptions of the trials have been previously reported (62, 63). The patients were recruited between 2015 to 2019. Details of surgical techniques and postoperative cares are included in the Supplementary Materials. Patient’s identifiable information was handled according to approved IRB protocol.

### Imaging Data

Patients underwent MR imaging of the knee at multiple time points between 3 to 24 months after ACL surgery. A single 3T scanner (Tim Trio, Siemens, Erlangan, Germany) and a 15-channel knee coil was used to scan the surgically treated and contralateral ACL-intact knees of each subject using common clinical sequences including Proton Density-weighted (PD-SPACE), T_2_-weighted Turbo Spin-Echo (T2-TSE) and Proton Density-weighted Turbo Spin-Echo (PD-TSE) along with a 3D Constructive Interference in Steady State (CISS) sequence (Table 1).

**Table 1.** MR imaging acquisition parameters and distribution.

| **Sequence Name** | **Acquisition Parameters** | **Number of Available MRI Sets** |
| --- | --- | --- |
| CISS | Sagittal Plane; TR/TE=14/7 msec, FA=35, 16cm FOV, 80x512x512 (slice x frequency x phase) | 677 |
| PD SPACE | Sagittal Plane, TR/TE = 1000/49 msec, 16 cm FOV, 0.5 mm slice thickness and 320 x 320 (phase x frequency) matrix | 405 |
| PD-TSE | Coronal Plane, TR/TE = 1900/27 msec, 14 cm FOV, 3 mm slice thickness and 384 x 384 (phase x frequency) matrix | 307 |
| T2-FS | Axial Plane: TR/TE = 4300/83 msec, 16 cm FOV, 1 mm slice/gap, 256 x 256 matrix | 373 |

The surgically treated ligaments and the contralateral native ACLs were manually segmented using image processing software (Mimics (v17.0; Materialize, Belgium)). The segmentation was done by an experienced investigator (AMK) with a high intra-rater reliability (ICC>0.9 for segmenting intact and surgically treated ACL) (60, 61). MR images were resampled to the isotropic voxel size (0.5mm x 0.5mm x 0.5mm), and then resized to 320 x 320 x 264 voxels. The manually segmented ACL masks were center-cropped into a size of 128 x 128 x 64 (Supplementary Fig S1). Each MRI was augmented (i.e., Gaussian noise, Gaussian blur, 3D rotation, and 3D translation) to increase the sample size and variability, and to generate a balanced distribution of all ACL types. The combination of raw and augmented MRI segmentations was then randomly divided into training and testing sets (Table 2). This split was stratified by subject to ensure that the subjects from the testing set were completely unseen to the model, and the model would not be evaluated by subjects from the training set.

**Table 2.**
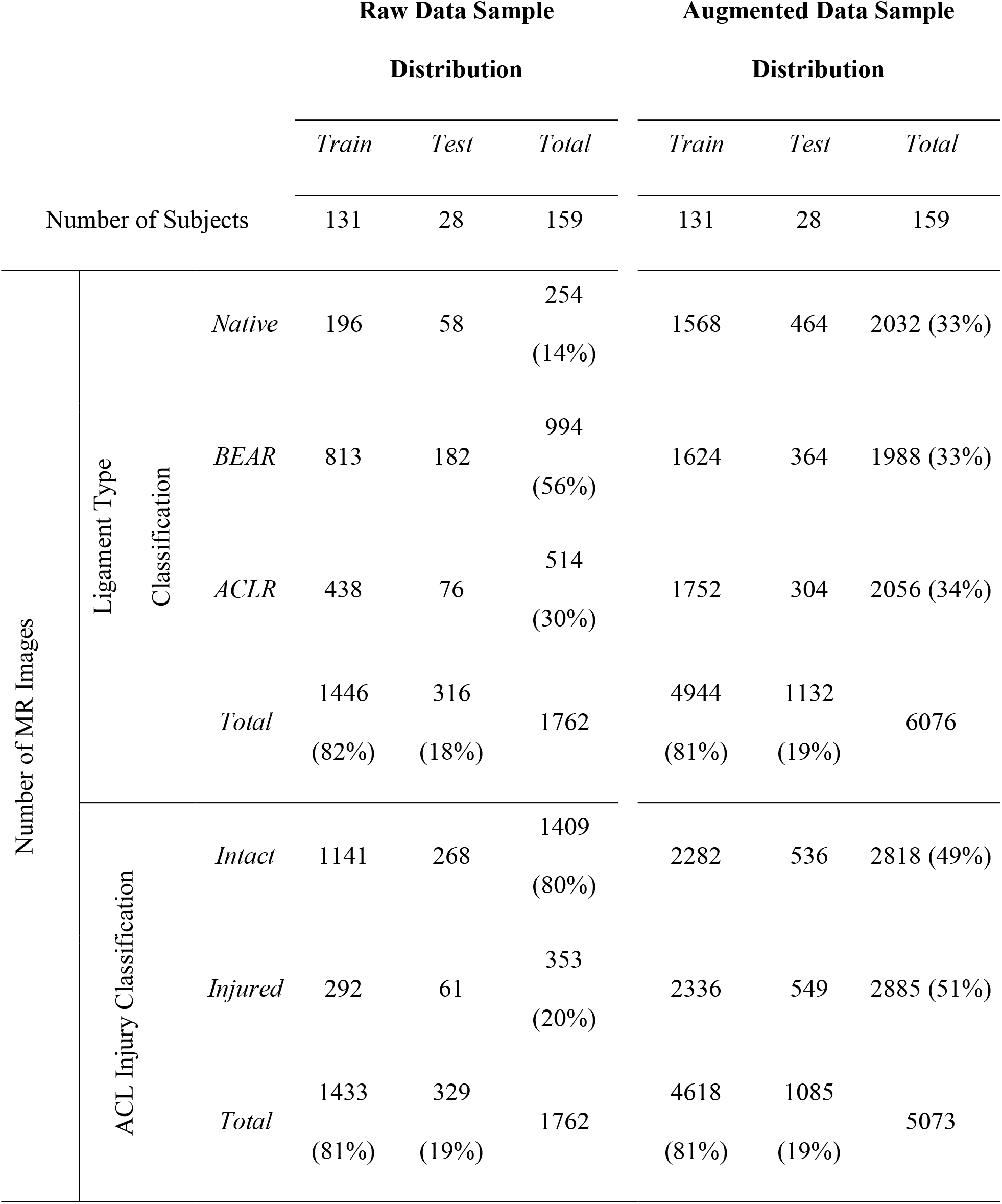
Distribution of subjects and imaging data between classes, and test and training sets.

### Non-Imaging Predictors

In addition to the MR images, we also collected 11 non-imaging demographics and clinical outcome measures including, age, sex, meniscus injury, time to return to sports (days), in addition to common clinical outcomes often studied in the context of ACL injury and recovery including patient reported outcomes (i.e. IKDC score) and functional assessments focused on knee laxity, muscle strength and single legged hop performance and knee range of motion (Supplementary Materials). The choice of these predictors was based on their clinical relevance and availability of data for all the corresponding MRIs. The collected non-imaging clinical data for each patient visit were represented with a vector of 11 elements. Hence, each MR image has a corresponding vector of clinical data. To address missing data points, we used imputation, which aims at inferring plausible values for the missing predictors (64). Based on preliminary cross-validation experiments, we chose a median-based imputation approach, which showed to result in higher classification accuracy compared to other imputers such as mean, constant value, and K nearest neighbors. The non-imaging clinical data were then normalized by removing the mean and scaling to unit variance, separately for each of the 11 variables. The non-imaging training dataset was up-sampled using SVM-SMOTE algorithm (65) to increase the sample size and to balance the data distribution over different classes.

### Ligament type classifier and feature extractor

To learn detailed features of the ACL and their differences between native and surgically treated tissues, we built and trained a 3D CNN feature extractor to discriminate the type of ligament (i.e., native, BEAR and ACLR) from the isolated MRI-segmented ACLs (Fig 1). We chose isolated segmented ACL as the input instead of the whole-knee MRI since the feature extractor was expected to focus on structural features of the ACL and not be distracted by other bold features of the surrounding tissue structures (e.g., bone tunnels in surgically treated ACLs). We built the CNN feature extractor based on a 3D Inception-ResNet architecture (66). It includes 9 Inception-ResNet blocks consisting of 3D convolutional layers. The CNN takes segmented ACL masks of size 128 x 128 x 64 voxels as input. The Inception-ResNet modules include multiple convolutional layers and residual connections that add the output of the convolutional layers to the input to ease the training of deep neural networks and increase the richness of the learned representations. Because of the large number of convolutional operations, 3D Inception-ResNet modules have a very large memory footprint, especially for large 3D images. To reduce the GPU memory requirements of our model, we used a series of 3D convolutional layers followed by max-pooling prior to the 3D Inception-ResNet modules in our network design. By this design, we reduced the size of the input to the Inception-ResNet blocks through feature learning. The size of the final encoded representation was 32 x 32 x 16 x 256, which was then fed into a global average pooling layer to further reduce its size. The output of global averaging passed through a softmax layer to obtain the conditional class probability estimates of the three classes of native, BEAR, and ACLR. The model was optimized to minimize the cross-entropy loss between the estimated and true class probability vectors. The optimization was performed using the Adam optimizer with a learning rate of 10^-5^.

**Fig 1.**
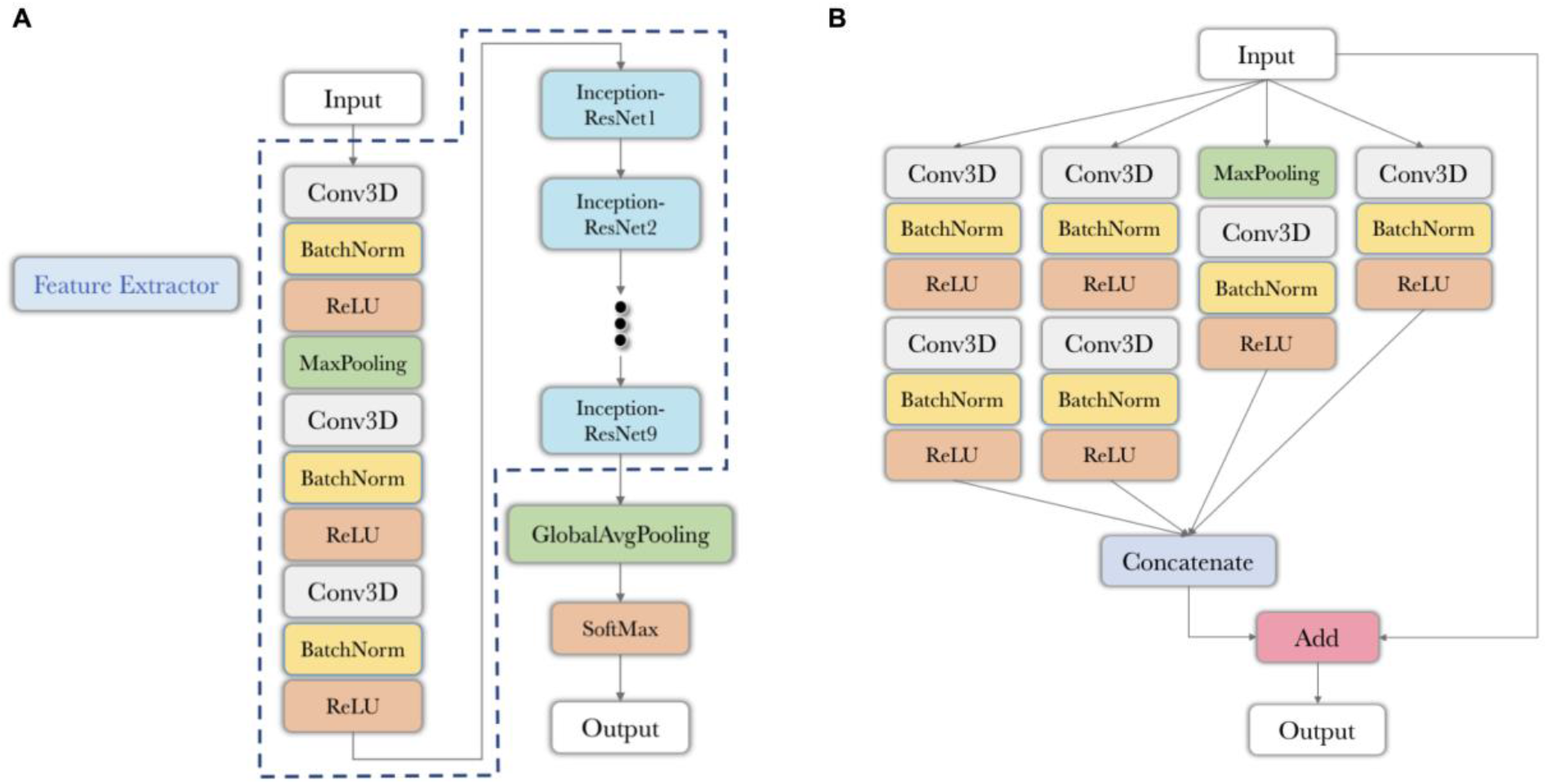
3D CNN feature extractor to classify ligament type from isolated MRI-segmented ACLs. (A) The structure of the 3D feature extractor. (B) The structure of 3D Inception-ResNet block. ReLU: rectified linear unit; Conv3D: 3D convolutional layer; BatchNorm: batch normalization; MaxPooling: max pooling layer; GlobalAvgPooling: global average pooling layer; SoftMax: softmax layer.

To assess the relative performance of the deep learning classier compared to human examiner, a randomly selected subset of 84 MRI-segmented ACLs from the testing set was reviewed by three experienced orthopedic surgeons involved in BEAR trials to independently identify the ACL type from ACL structure alone (Supplementary Fig S2). This was done through an online survey (Qualtrics XM, Qualtrics, Seattle, WA). All surgeons classified the same ACLs, but the orders were randomized between the examiners. The performance metrics for all the three examiners are presented in.

### ACL injury classifier from the isolated ACL MRI-segmentations

The trained classifier described above was used as a base feature extractor in building a model for subsequent ACL injury risk prediction. This was done by adding several new layers on top of the feature extractor network (Fig 2), where the feature extractor structure is shown in Fig. 1A. These additional layers consist of a standard 3D convolutional layer followed by an additional Inception-ResNet block. The final part of this model is a softmax layer that outputs a binary probability vector for Intact versus Injured ACL classification. The predicted probability of the injured ACL class is identified as the injury risk score. The injury label was defined based on the ACL-related adverse events with MRI and non-imaging data preceding any ACL-related adverse event (e.g., instability, confirmed ACL tear). To train this new model, the layers that were previously trained as part of the feature extractor network training were kept fixed. Only the new top layers were trained based on the binary labels of intact vs injured. Given the class imbalance between intact and injured classes, rather than using the cross-entropy loss (used in the feature extractor), in this step, we used the Tversky loss for training. This loss has been shown to be more effective for applications where the data samples with positive labels account for a small fraction of the available training data.

**Fig 2.**
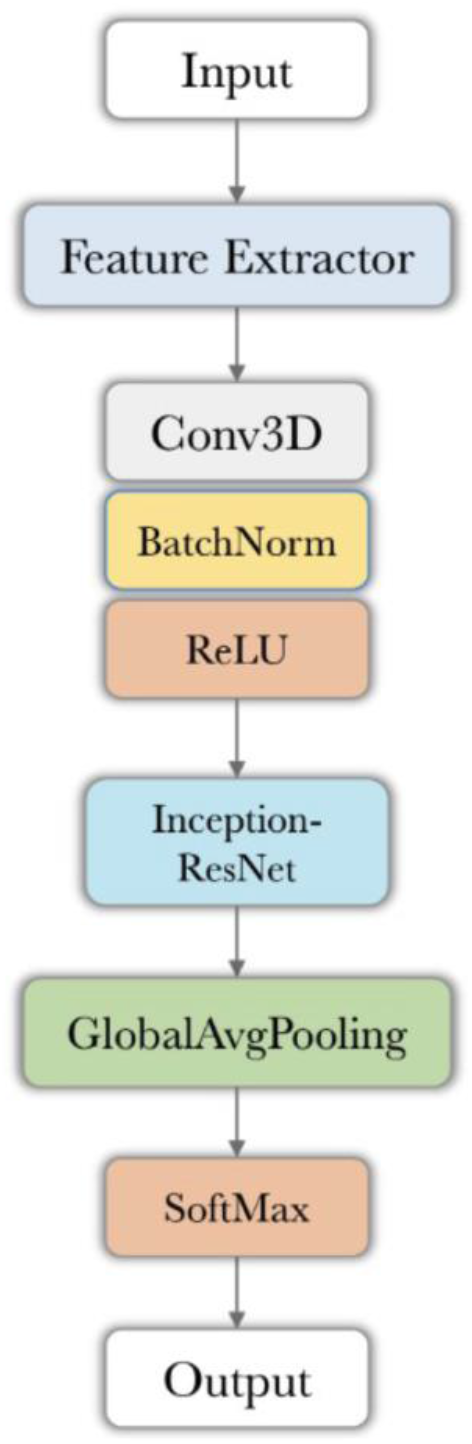
ACL injury classifier from isolated ACL MRI segmentations. The structures of Feature Extractor and Inception-ResNet block are shown in Fig 1A and 1B, respectively.

### ACL injury classifier from whole-knee MRI

We also train a separate CNN model to exploit the features in the whole-knee MRI to predict subsequent ACL injury. This model takes a 3D knee MRI image as input and outputs an ACL injury risk score, which is the probability of the input image belonging to the injured ACL class. The architecture of this CNN model was identical to that of the feature extractor CNN described above. To adapt that architecture to this new setting, we modified the sizes of the input and output layers in order to accommodate the full knee MRI as input and output a two-class probability vector. Instead of training the CNN parameters from scratch, we initialized them with the feature extractor model weights and then fine-tuned the entire model for the new task using the Tversky loss function. For this classifier, the native ACLs were removed as they did not have any surgery related landmarks (e.g., bone tunnels) which is highly likely to be precepted by the CNN as a feature of ACL injury, and thus may lead to biased predictions.

### ACL injury classifier from non-imaging predictors

We aimed to investigate if the integration of relevant non-imaging clinical data improves the risk prediction of subsequent ACL injury. To implement this pipeline, we first developed a non-imaging-based classifier based on 11 demographics and clinical outcome measures that are commonly recorded during postoperative visits and have shown to be important factors related to ACL function and lower extremity biomechanics. Using a logistic regression classifier and an 11-dimensional non-imaging input vector, we predict the probability of the Injured ACL class as the ACL injury risk score. The choice of logistic regression was based on its superior performance compared to other classifiers as shown in Supplementary Fig S3. The LBFGS optimizer approximates the second derivative matrix updates with gradient evaluations during the back-propagation optimization (67).

### Probability fusion in the multi-modal pipeline

The final module in our proposed machine learning pipeline is a fusion module to integrate the ACL injury risks estimated by classifiers described above and output a final estimate of the risk of injury (Fig 3). We anticipate that the risk estimated by the fusion module would be more accurate than those estimated by each of the three modules independently, since the fusion module synergistically integrates the information extracted from multiple data. In order to investigate the importance of different sources of information in the risk estimation problem, we experimented with three different implementations of the fusion model: The first fusion implementation used only risk scores from isolated ACL MRI segmentation CNN and the non-imaging classifier. It ignores the risk estimated by Module B from the whole-knee MRI CNN. The second implementation uses only risk score from whole-knee MRI CNN and the non-imaging classifier. The third implementation uses the risk estimated by all three modules as input. The risk score distributions estimated based on different modalities were concatenated into a single matrix and then fed into the fusion model. The output of the model is a single estimated risk score for subsequent ACL injury merged from different data sources. The fusion models for different modality combinations were all performed by Logistic Regression classifier, using the same LBFGS algorithm minimizing the cross-entropy loss. The choice of Logistic Regression to fuse the risk scores is based on its performance compared to other approaches to distinguish between intact and injured cases (Supplementary Fig S4).

**Fig 3.**
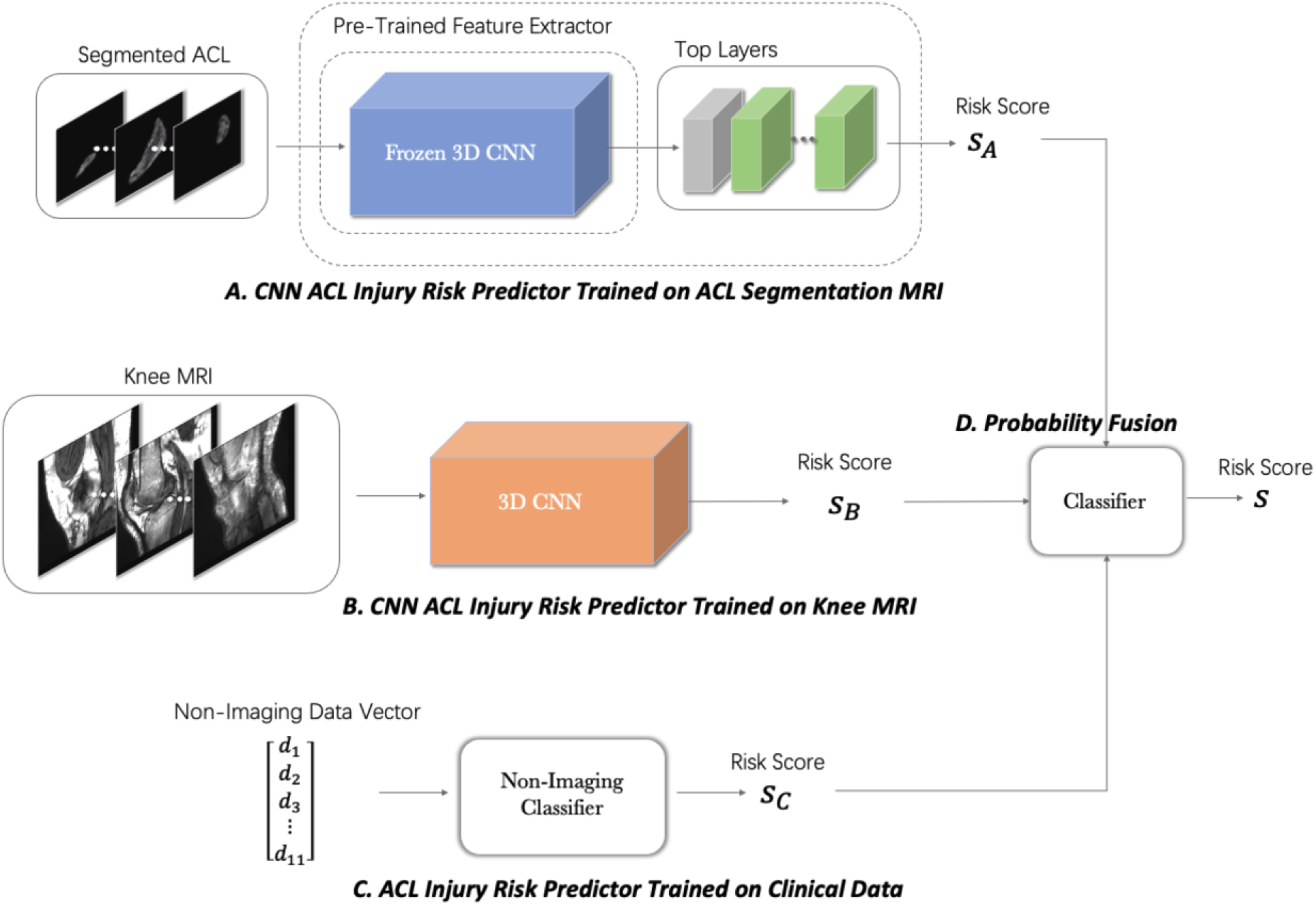
Multi-modal pipeline to predict the risk of subsequent ACL injury based on a combination of the imaging and non-imaging predictors. The model classifies the ACL to intact or injured by fusing the probability of injury (risk score) from isolated MRI-segmented ACL (A), whole-knee MRI (B) and non-imaging data (C) using a logistic regression classifier (D).

### Statistical Analysis

Performance measures for the models and orthopedic surgeons included sensitivity, specificity, and accuracy. We also evaluated the trade-off between the true positive rates and the positive predictive values for all models using precision-recall curves. We assessed the model’s performance with the area under the receiver operating characteristic curve (AUROC) and the area under the precision-recall curve (AUPRC). To assess the variability in estimates, we provide 95% confidence intervals for sensitivity, specificity, accuracy, AUROC and AUPRC. DeLong test (68) as used to compare the AUROC between different injury classifiers. Bonferroni posthoc was used to correct the p values from multiple comparisons. All statistical analyses were completed in the R environment for statistical computing.

## RESULTS

### Ligament type classifier and feature extractor

The trained model achieved an ACL type classification accuracy of 93.2% (91.7% – 94.7%), area under the receiver operating characteristic curve (AUROC) of 0.99 (0.98 – 0.99), and area under the precision-recall curve (AUPRC) of 0.98 (0.97 – 0.99) on the testing set (Fig 4). The trained feature extractor was able to identify type of ACL with 94.6% sensitivity and 96.0% specificity. The human examiners had a substantially inferior performance (accuracy: 0.45 – 0.48, AUROC: 0.54 – 0.66, AUPRC: 0.25 – 0.60) compared to CNN feature extractor on all quantified performance metrics (Supplementary Fig S5).

**Fig. 4.**
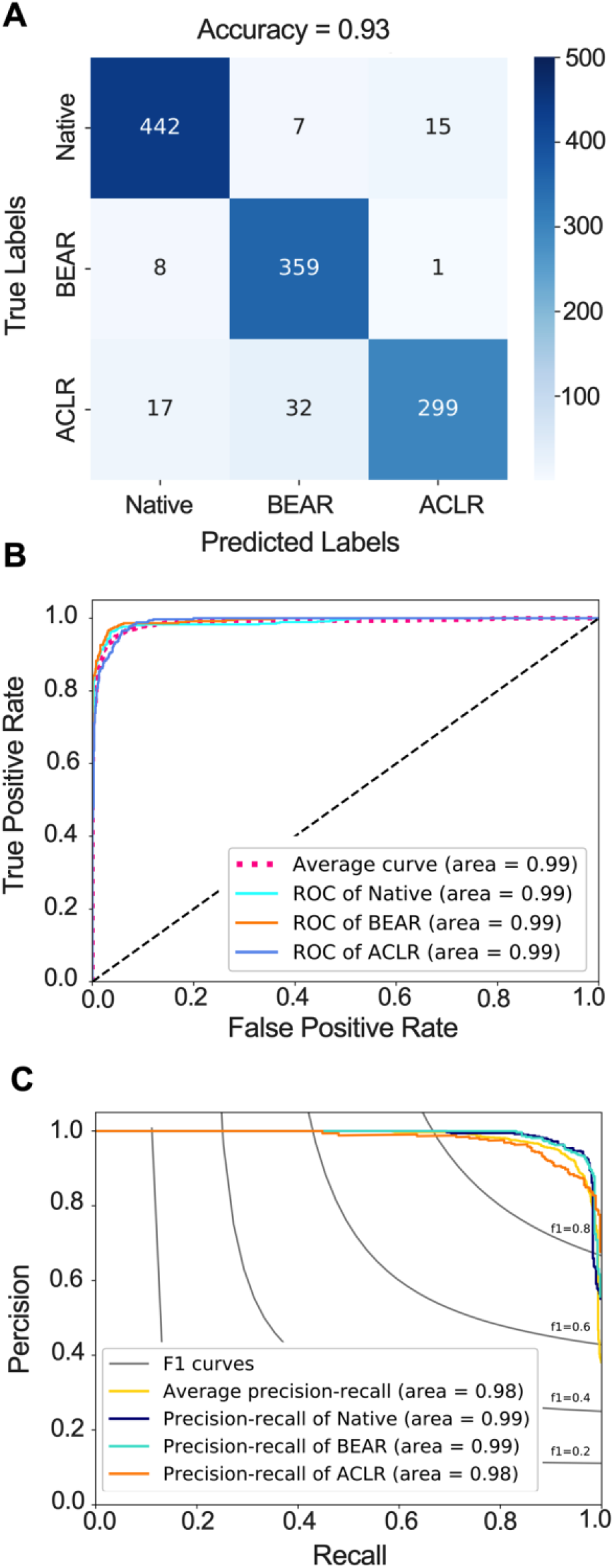
Feature extractor performance in classifying ACL type from isolated MRI-segmented ACL. **(A)** Confusion matrix. **(B)** Receiver operating characteristic (ROC) curve. **(C)** Precision recall curve. BEAR: Bridge-Enhanced ACL Restoration, ACLR: ACL Reconstruction.

### ACL injury classifier from the isolated ACL MRI-segmentations

The trained ACL injury classifier achieved a classification accuracy of 77.6%, (75.2% – 80.1%), AUROC value of 0.84 (0.82 – 0.86), and AUPRC values of 0.84 (0.81 – 0.87) on the testing set (Fig 5A-C). The trained CNN was able to predict ACL injury with 75.3% sensitivity and 80.6% specificity.

**Fig 5.**
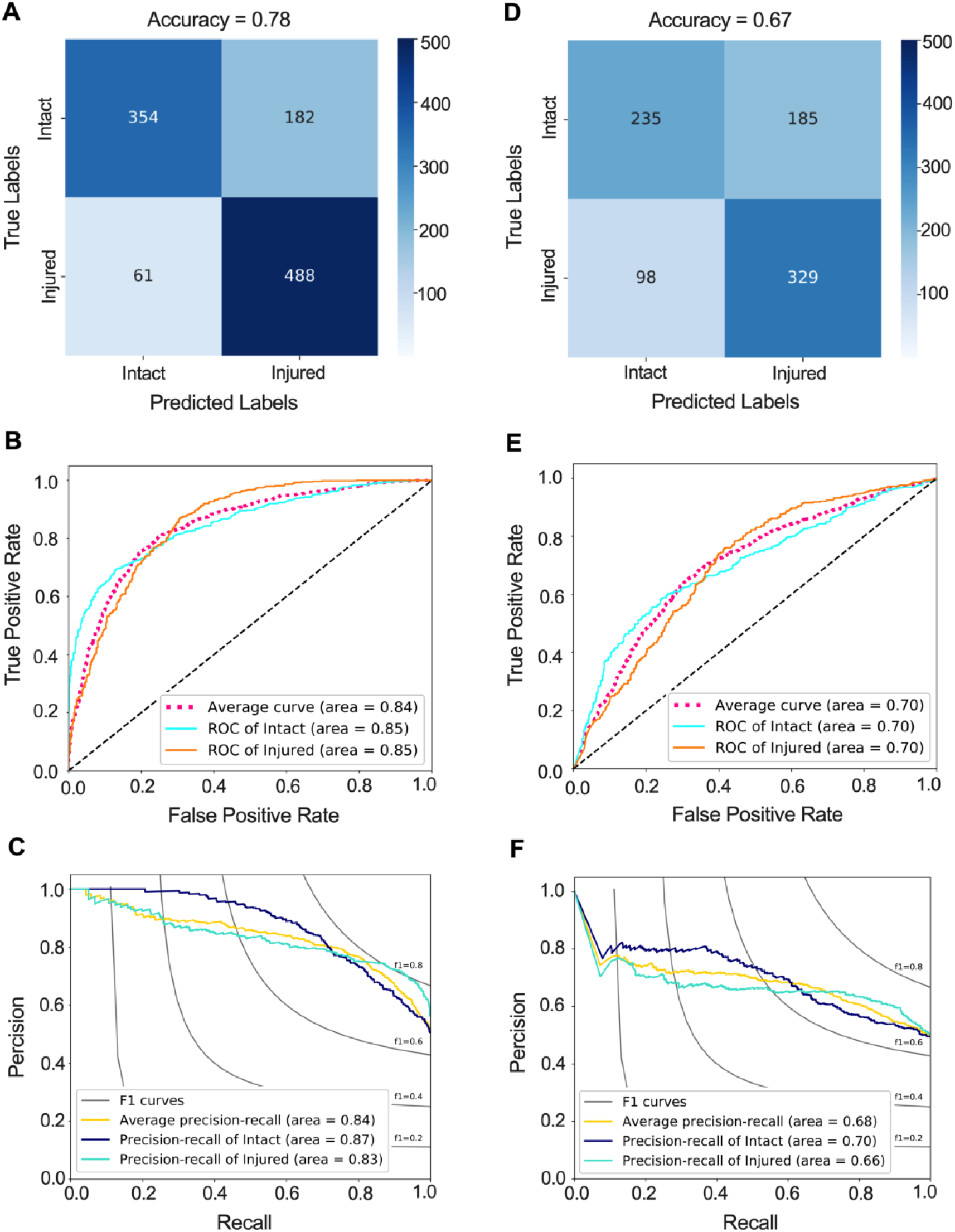
CNN classifier performance in predicting the subsequent ACL injury. **(A)** Confusion matrix, **(B)** receiver operating characteristic (ROC) curve, and **(C)** precision recall curve for CNN classifier that predicts the subsequent ACL injury based on isolated MRI-segmented ACLs. **(D)** Confusion matrix, **(E)** ROC curve, and **(F)** precision recall curve for CNN classifier that predicts the subsequent ACL injury based on whole-knee MRIs.

### ACL injury classifier from whole-knee MRI

The model resulted in classification accuracy of 66.6% (63.8% – 69.4%), AUROC value of 0.70 (0.67 – 0.73) and AUPRC value of 0.68 (0.64 – 0.72) on independent testing set (Fig 5D-F). The trained CNN was able to predict ACL injury with 68.7% sensitivity and 65.4% specificity. The AUROC for the whole-knee MRI classifier was significantly smaller than the AUROC for isolated MRI-segmented ACL classifier (Adjusted P<0.001).

### ACL injury classifier from non-imaging predictors

The model achieved a classification accuracy of 70.1% (67.4% – 72.8%), AUROC value of 0.75 (0.72 – 0.78) and AUPRC value of 0.72 (0.69 – 0.76) on the testing set (Fig 6). The trained model was able to predict ACL injury with 69.0% sensitivity and 71.4% specificity. While the AUROC for the non-imaging classifier was not significantly different from the AUROC of the whole-knee MRI classifier (Adjusted P = 0.210), it was significantly lower than the AUROC of the isolated MRI-segmented ACL classifier using (Adjusted P = 0.039).

**Fig 6.**
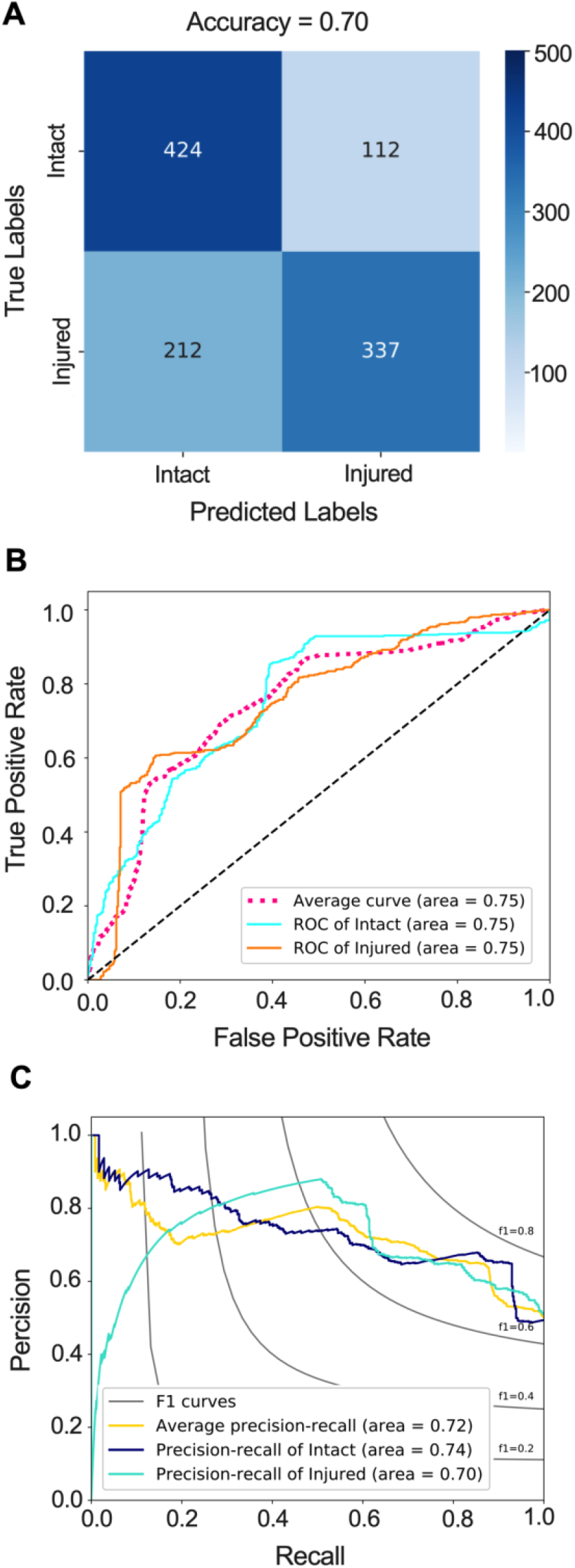
Logistic regression model performance in predicting subsequent ACL injury from non-imaging predictors. **(A)** Confusion matrix. **(B)** Receiver operating characteristic (ROC) curve. **(C)** Precision recall curve.

### Multi-modal pipeline to predict subsequent ACL injury

In order to investigate the importance of different sources of information and to find the best multi-modal pipeline structure to predict subsequent injury risk, we evaluated three different fusion strategies. In the first model, we fused the probability of injury risk obtained from the ACL segmentations CNN classifier with those from non-imaging classifier. This model yielded classification accuracy of 79.9% (77.5% – 82.3%), AUROC value of 0.88 (0.86 – 0.90) and AUPRC value of 0.88 (0.85 – 0.90) on the testing set (Fig 7A-C). The fusion of isolated MRI-segmented ACL CNN and non-imaging classifiers was able to predict injury risk with 80.0% sensitivity and 80.4% specificity. This fusion of these two classifiers resulted in a higher AUROC compared to the AUROC of isolated MRI-segmented ACL classifier (Adjusted P<0.001) and AUROC of non-imaging classifier (Adjusted P<0.001).

**Fig 7.**
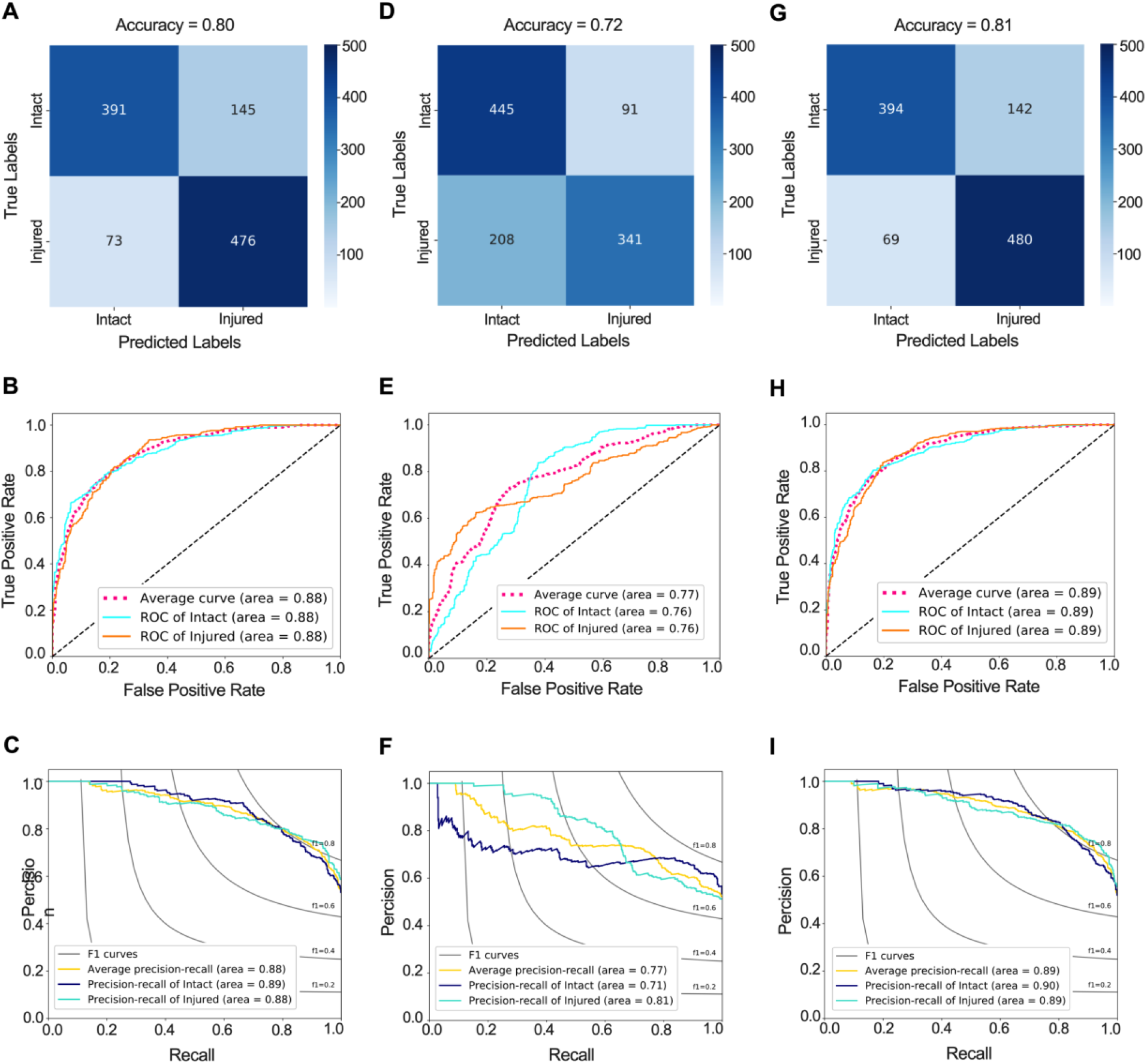
Multi-modal classifier performance in predicting the subsequent ACL injury. **(A)** Confusion matrix, **(B)** receiver operating characteristic (ROC) curve, and **(C)** precision recall curve for multi-modal classifier that predicts the subsequent ACL injury based on isolated MRI-segmented ACL and non-imaging predictors. **(D)** Confusion matrix, **(E)** ROC curve, and **(F)** precision recall curve for multi-modal classifier that predicts the subsequent ACL injury the risk of injury based on whole-knee MRI and non-imaging predictors. **(G)** Confusion matrix, **(H)** ROC curve, and **(I)** precision recall curve for multi-modal classifier that predicts the subsequent ACL injury based on isolated MRI-segmented ACL, whole-knee MRI and non-imaging predictors.

The second fusion strategy included the whole-knee MRI CNN and non-imaging classifiers which resulted in classification accuracy of 72.4% (69.7% – 75.1%), AUROC value of 0.77 (0.74 – 0.80) and AUPRC value of 0.77 (0.74 – 0.81) on the testing set (Fig 7D-F). This model was able to predict subsequent injury risk with 72.4% sensitivity and 72.5% specificity. There were no differences in AUROC between this fused classifier, and individual whole-knee MRI and non-imaging classifiers (Adjusted P>0.2 for all comparisons). However, the AUROC of this classifier was significantly lower than the AUROC of the classifier based on the fusion of the ACL segmentation and non-imaging clinical predictors (Adjusted P<0.001).

The fusion of all three classifiers resulted in highest classification accuracy (80.6% (78.2% – 83.0%)), AUROC value (0.89 (0.87 – 0.91)) and AUPRC value (0.89 (0.86 – 0.92)) and was able to predict subsequent injury risk with 77.2% sensitivity and 85.6% specificity (Fig. 7G-I). This multi-modal classifier had a higher AUROC compared to each individual classifier and the classifier based on the fusion of the whole-knee MRI with non-imaging predictors (Adjusted P<0.001 for all comparisons). There was no significant difference in AUROC of this multi-modal classifier and the classifier based on the fusion of the ACL segmentation and non-imaging clinical predictors (Adjusted P=0.801).

### Visualization of ACL features and injury biomarkers

To visualize the MRI features contributing to model’s prediction subsequent ACL injury risk, occlusion maps (54) were generated for true positive cases identified by the isolated MRI-segmented ACL CNN classifier. A representative set of these images from the central slice of the ACL are shown in Fig 8. As seen in these examples, the hot zone of the map, indicating the most important features contributing to model classification decision of injury, is consistently within the distal half of the ACL. This pattern is even consistent across different MRI sequences of the same subjects, highlighting the model’s ability to evaluate injury risk from ACL patterns independent of MRI sequence. This is further supported by minimal differences in classifiers’ performance using each individual MRI sequence (Fig 9). There were no significant differences in the AUROC of the subsequent ACL injury risk classifiers utilizing different MRI sequences (Adjusted P>0.1 for all comparisons).

**Fig 8.**
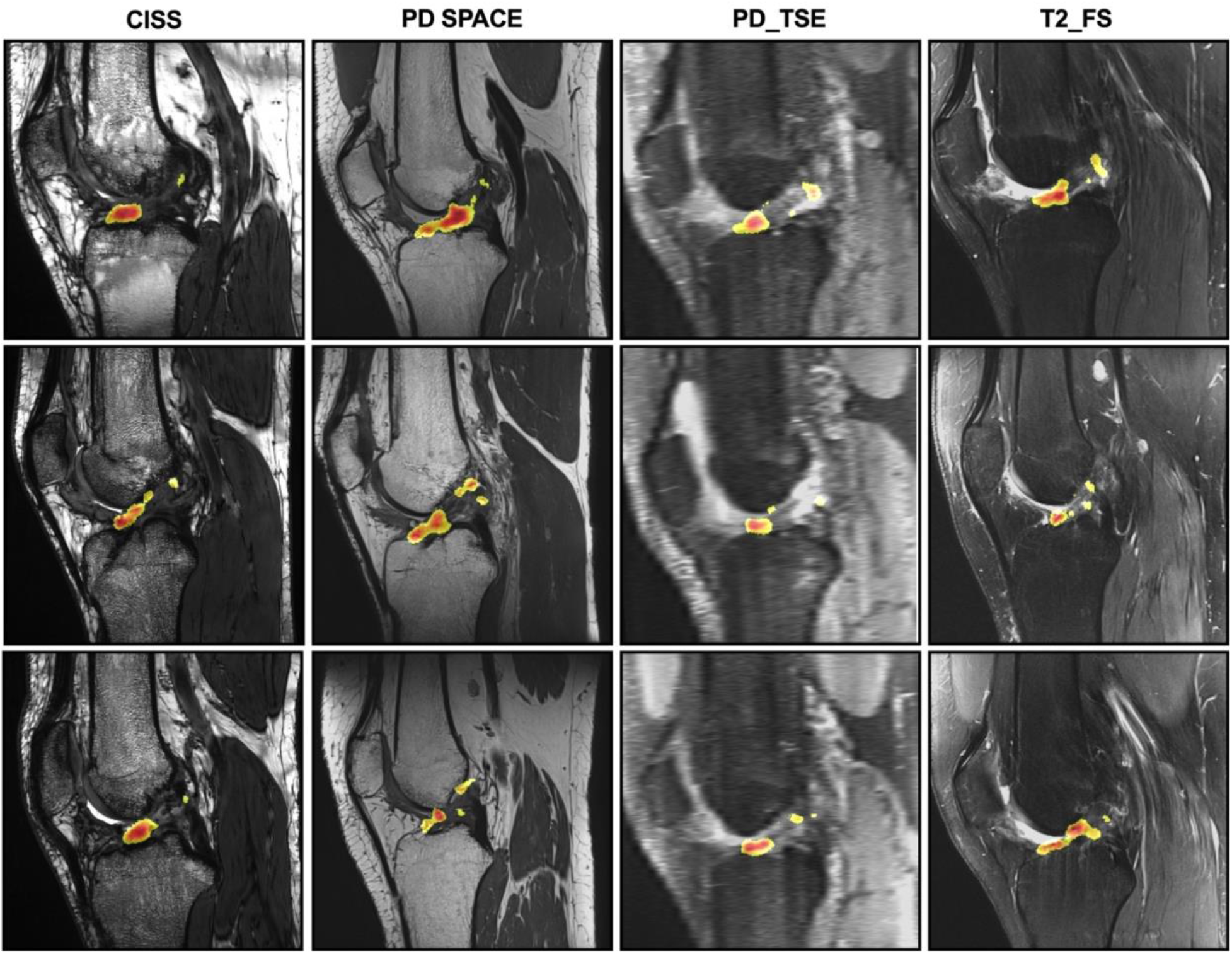
Representative central slice view of the occlusion maps superimposed on their corresponding MRI slice. The maps were generated for all the true positive test cases (knees with subsequent ACL injury). Each column includes the representative maps from true positive cases of each MRI sequence. Each row shows the occlusion maps for the same knee under different MRI sequences. The occlusion maps are generated based on the isolated MRI-segmented ACL CNN classifier to identify ACL features contributing to correct decision by the classifier. The color map corresponds to the relative positive contribution of the pixel in classifiers ability to make a correct assignment. The red color corresponds to highest positive contribution. The heatmap is generated on a scale of 0 (dark blue) to 255 (dark red). For better visibility, we only showed the top 50% of the heatmap (127 – 255). CISS: constructive interference in steady state, PD: proton density, TSE: turbo spin echo, FS: fat suppression.

**Fig 9.**
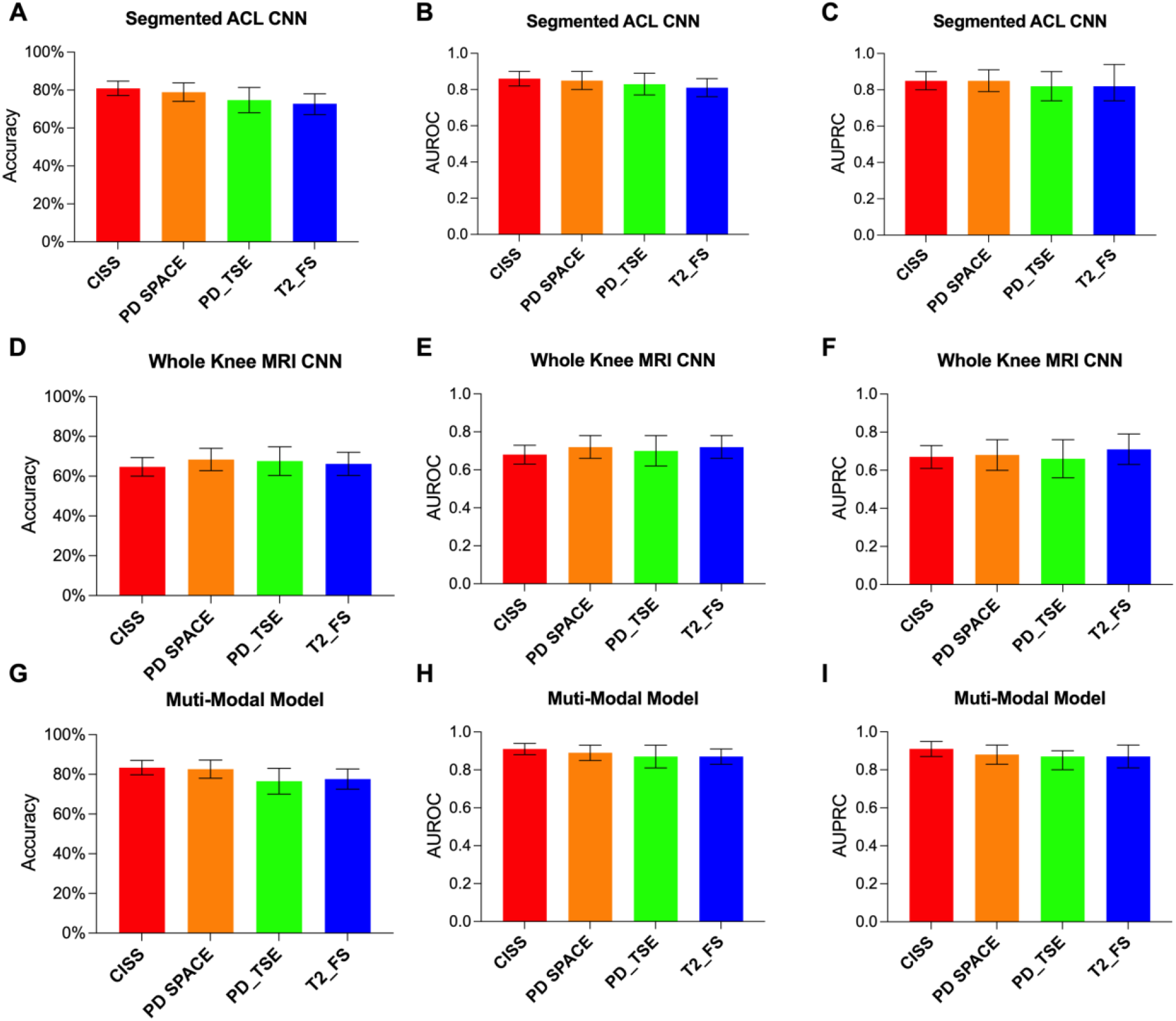
Classifiers’ performance metrics for each independent tested MRI sequence. Performance metrics in predicted risk of subsequent ACL injury for **(A-C)** CNN based on isolated MRI-segmented ACL, **(D-F)** CNN based on whole-knee MRI, and **(G-I)** Multi-modal model using isolated MRI-segmented ACL, whole-knee MRI and non-imaging clinical predictors. The values are presented as mean and the error bars represent the 95% CI. For all metrics, the confidence intervals are overlapping, indicating no significant differences between sequences. CISS: constructive interference in steady state, PD: proton density, TSE: turbo spin echo, FS: fat suppression, AUROC: area under the receiver operating characteristic curve, AUPRC: area under the precision-recall curve.

## DISCUSSION

Our results demonstrate that a deep learning approach can achieve high performance in identifying patients at high risk of subsequent ACL injury after ACL surgery. The data suggest that the deep learning pipeline has a superior performance in extracting ACL features required to differentiate between native ACLs, restored ACLs (BEAR) and reconstructed grafts (ACLR) compared to experienced clinicians. Results also show that building and applying the model to isolated MRI-segmented ACLs are better predictors of a subsequent ACL surgery than whole-knee MRIs and non-imaging clinical predictors; however, even better performance can be achieved by using a combination of isolated MRI-segmented ACLs, whole-knee MRI and non-imaging clinical predictors. Finally, our findings indicate that the proposed deep learning pipeline can classify the injured ACLs relatively independent of MRI sequence and have the capability to identify new imaging biomarkers to predict the risk of subsequent ACL injury following surgical treatment.

Qualitative assessment of knee MRI has been routinely used to confirm ACL or graft injury based on tissue appearance, including signal distribution and physical organization. Over the past two decades, several groups, including ours, have worked on developing quantitative MRI sequences and analysis techniques to systematically track healing ACL. These include measurements of signal intensity and T_2_* relaxometry to predict ACL structural properties in preclinical models (26-34) and to assess ACL remodeling after surgery in human patients(35-48). Recent studies have shown that deep learning-based analysis of knee MRIs can accurately classify type of soft tissue injury (i.e. ACL or meniscus tears)(59) and has been extensively used to assess knee osteoarthritis risk and progression (58, 69-71). These promising studies encouraged us to leverage the power of deep learning to improve upon previous ACL MRI studies to address a pressing unmet need in predicting the risk of subsequent ACL injury following surgical treatment.

In contrast to prior deep learning studies which use whole-knee MRI as the input to the network, here we started by training a feature extractor using isolated MRI-segmented ACL to focus on the tissue of interest. The feature extractor was able to identify ACL type (i.e., native, BEAR, ACLR) with an accuracy almost double that achieved by experienced human examiners. Using this feature extractor, we trained a classifier CNN which predicted the risk of subsequent ACL injury after surgery with superior accuracy, sensitivity and specificity compared to CNN classifiers based on whole-knee MRI or non-imaging clinical predictors. While the non-imaging classifier also had a higher accuracy, sensitivity, and specificity than the whole-knee MRI classifier, it had a less stable precision-recall curve when classifying injured cases. Interestingly, CNN classifiers based on the isolated MRI-segmented ACL or whole-knee MRI had higher rates of false positives than false negative, whereas the non-imaging CNN classifier resulted in lower false positives than false negatives. The fusion of MRI (i.e., isolated MRI-segmented ACL or whole knee) classifier predictions with non-imaging classifier predictions improved the model accuracy, sensitivity, and specificity by up to 7%, but most importantly, reduced the false positive rates in both classifiers. Additionally, fused models had more robust precision-recall curves than the non-imaging classifier. Ultimately, the multi-modal classifier, which leveraged the inputs from all three sources, achieved the highest performance in predicting subsequent ACL injury risk. While the multi-modal classifier had substantially better performance compared to whole-knee MRI classifier or non-imaging classifier (up to 10% improvement in performance metrics), it also had slightly better accuracy, sensitivity, and specificity compared to isolated MRI-segmented ACL classifier (<5% improvements). In addition, the multi-modal classifier resulted in ∼25% reduction in false positive rate compared to isolated MRI-segmented ACL classifier. These observations suggest that while a CNN classifier based on only the isolated MRI-segmented ACL could predict the risk of subsequent ACL injury, addition of non-ACL features (i.e., whole-knee MRI and non-imaging clinical data) improved the prediction specificity in particular with regards to false positives. The similarities in model performance and occlusion maps between the sequences are also very reassuring and suggest that such a deep learning pipeline can predict the risk of subsequent ACL injury based on a range of MRI sequences. This is an important advantage as a major shortcoming of current quantitative MRI assessments is their strong dependency on sequence and acquisition parameters, which limit their generalizability and clinical translation. The current proof-of-concept study suggests that the proposed platform may be able to address some of those shortcomings, although future multisite studies with data from different MRI magnets are required.

This study has limitations. Our training and test data are from a single site and single magnet, which limits the generalizability of our findings. Unfortunately, there are very few studies with postoperative ACL MRIs, long-term follow-up assessments, and adequate sample size to develop this model. However, data from the BEAR trials provided a range of relevant outcomes along with longitudinal postoperative MR images in large cohort of ACL injured patients to construct the models. The lack of similar datasets with available postoperative MRIs and follow up limited our ability to further evaluate our pipelines generalizability and prohibited us from external validation. This is primarily because the available literature and quantitative MRI techniques to assess healing ACL have not yet had an impact clinical care. As a result, postoperative MRI in ACL surgeries are not routinely performed. The findings reported in current study support the utility of postoperative MRI to predict the risk of subsequent ACL injury after surgery. We hope these results encourage more investigators to consider postoperative MRI in large scale ACL cohorts. Another limitation is the inclusion of different surgical treatments (BEAR and ACLR), which may have negatively influenced the model’s performance. Future studies with larger numbers of patients across each treatment will enable us to optimize the model performance based on treatment strategy. We have taken every possible measure (e.g., randomization, blinding, quality control) to minimize the bias and errors related to these limitations. Altogether, these findings should be considered as proof-of-concept, they highlight the potential of this computational deep learning approach in impacting the clinical care, which requires further research, improvement and validation.

Recent advances in deep learning along with improved understanding of the imaging markers and their links to tissue healing have paved the way for the development of advanced deep learning platforms to transform the clinical care of patients with traumatic joint injuries, such ACL tears. The work presented here is an example of how a multi-modal deep learning pipeline can be used to predict the risk of subsequent ligament injury following surgical treatment. The current study justifies further development of such a platform with a more robust, generalized, and validated performance. Upon successful development and translation, such an approach could be used to assist clinical care teams to better manage the postoperative care of patients with ACL injuries and has the potential to be transferred to evaluate other soft tissue disorders in a more data-driven and personalized manner.

## Data Availability

The clinical and imaging data for this study are form a series of FDA-regulated clinical trials (NCT02292004, NCT02664545, NCT03348995), which cannot be publicly shared due to privacy and regulatory issues. These trails have been extensively published. All the necessary details, required to replicate the studies here are included in the main text and supplementary materials. Additional data and models can be requested following proper materials transfer agreements.

## Acknowledgments

We would like to acknowledge the significant contributions of the clinical trial team including Bethany Trainor. We would also like to acknowledge the contributions of our medical safety monitoring team of Joseph DeAngelis, Peter Nigrovic, and Carolyn Hettrich, our data monitors Maggie Malsch, Meghan Fitzgerald, and Erica Denhoff, as well as the clinical care team for the trial patients, including Kathryn Ackerman, Alyssa Aguiar, Judd Allen, Michael Beasley, Jennifer Beck, Dennis Borg, Jeff Brodeur, Stephanie Burgess, Melissa Christino, Sarah Collins, Gianmichel Corrado, Sara Carpenito, Corey Dawkins, Pierre D’Hemecourt, Jon Ferguson, Michele Flannery, Casey Gavin, Ellen Geminiani, Stacey Gigante, Annie Griffin, Emily Hanson, Elspeth Hart, Jackie Hastings, Pamela Horne-Goffigan, Christine Gonzalez, Meghan Keating, Elizabeth KillKelly, Elizabeth Kramer, Pamela Lang, Hayley Lough, Chaimae Martin, Michael McClincy, William Meehan, Ariana Moccia, Jen Morse, Mariah Mullen, Stacey Murphy, Emily Niu, Michael O’Brien, Nikolas Paschos, Katrina Plavetsky, Bridget Quinn, Shannon Savage, Edward Schleyer, Benjamin Shore, Cynthia Stein, Andrea Stracciolini, Dai Sugimoto, Dylan Taylor, Ashleigh Thorogood, Kevin Wenner, Brianna Quintiliani, and Natasha Trentacosta. We would also like to thank the perioperative and operating room staff and the members of the Department of Anesthesia who were extremely helpful in developing the perioperative and intraoperative protocols. We would also like to acknowledge the efforts of other scaffold manufacturing team members, including Gabe Perrone, Gordon Roberts, Doris Peterkin, and Jakob Sieker. We are also grateful for the study design guidance provided by the Division of Orthopedic Devices at the Center for Devices and Radiological Health at the U.S. Food and Drug Administration under the guidance of Laurence Coyne and Mark Melkerson, particularly the efforts of Casey Hanley, Peter Hudson, Jemin Dedania, Pooja Panigrahi, and Neil Barkin. We are also especially grateful to the patients and their families who participated in this study, their willingness to participate in research that may help others in the future inspires all of us. We would like to acknowledge funding support from:

Boston Children’s Hospital Research Faculty Council (AMK)

Children’s Hospital Orthopaedic Surgery Foundation (MMM and AMK)

Children’s Hospital Sports Medicine Foundation (MMM)

Translational Research Program at Boston Children’s Hospital (MMM)

NIH/NIAMS R01-AR065462 (MMM, BCF and AMK)

NIH/NIAMS R01-AR056834 (MMM and BCF)

Lucy Lippitt Endowment of Brown University (BCF)

Rhode Island Hospital Orthopaedic Surgery Foundation (BCF)

NIH/NIGMS P20-GM103645 and P30-GM122732 (BCF)

Football Players Health Study at Harvard University (MMM and AMK). The Football Players Health Study is funded by a grant from the National Football League Players Association. The content is solely the responsibility of the authors and does not necessarily represent the official views of Harvard Medical School, Harvard University or its affiliated academic health care centers, the National Football League Players Association, Boston Children’s Hospital or the National Institutes of Health.

## Author contributions

Conceptualization: AMK, MMM, BCF

Methodology: AMK, AG, BCF, SWF, MH, DK, MS, JYK, BEAR Trial Team, KE, MMM

Investigation: AMK, AG, BCF, SWF, MH, DK, MS, JYK, BEAR Trial Team, KE, MMM

Visualization: AMK, AG, DK, MH, MS, IYK, BCF

Funding acquisition: AMK, MMM, BCF

Project administration: AMK, MMM, BCF

Supervision: AMK, BCF, MMM, AG

Writing – original draft: AMK, MH, MS, AG, DK

Writing – review & editing: All the co-authors

