## Supplementary Materials for "LigaNET: A multi-modal deep learning approach to predict the risk of subsequent anterior cruciate ligament injury after surgery"

Materials and Methods

### ***ACL reconstruction with autograft tendon (ACLR)***

### A standard hamstring autograft procedure was performed using a quadruple semitendinosus-gracilis graft (n=33) or central third bone-patellar tendon-bone autograft (n=2) using a continuous-loop cortical button (Endobutton; Smith & Nephew, Andover, MA) for proximal fixation and a bioabsorbable interference screw (BioRCI HA; Smith & Nephew) for tibial fixation. A minimal notchplasty was performed at the surgeon’s discretion as needed for adequate visualization of the posterior notch for placement of the femoral tunnel starting point within the prior ACL footprint. The femoral tunnel was drilled using an anteromedial portal technique and a flexible drill system (Clancy Anatomic Cruciate Guide, Smith & Nephew).

### ***Bridge-Enhanced ACL Restoration (BEAR)***

### After the induction of general anesthesia, an examination was performed to verify the positive pivot shift on the injured side and to record the Lachman test, range of motion and pivot shift exam results on both knees. A tourniquet was then applied to the surgical limb. A knee arthroscopy was performed, and any meniscal injuries were treated if present. A tibial aimer (ACUFEX Director Drill Guide; Smith and Nephew, Andover, MA) was used to place a 2.4mm guide pin through the tibia and the tibial footprint of the ACL. The pin was over-drilled with a 4.5 mm reamer (Endoscopic Drill; Smith & Nephew, Andover, MA). A notchplasty was performed using a combination of shaver and curette to facilitate visualization of the femoral footprint. A guide pin was then placed in the femoral ACL footprint, drilled through the femur and then over-drilled with the 4.5 mm reamer. A 4 cm arthrotomy was made at the medial border of the patellar tendon and a whip stitch of #2 absorbable braided suture (Vicryl; Ethicon, Cincinnati, OH) was placed into the tibial stump of the torn ACL. Two #2 non-absorbable braided sutures (Ethibond; Ethicon, Cincinnati OH) were looped through the two center holes of a cortical button (Endobutton; Smith & Nephew, Andover, MA). The free ends of a #2 absorbable braided suture from the tibial stump were passed through the cortical button, which was then passed through the femoral tunnel and engaged on the lateral femoral cortex. Both looped sutures of #2 non-absorbable braided (four matched ends) were passed through the scaffold, and 10 cc of autologous blood obtained from the antecubital vein was added to the scaffold. The scaffold was then passed up along the sutures into the femoral notch and the non-absorbable braided sutures were passed through the tibial tunnel and tied over a second cortical button on the anterior tibial cortex with the knee in full extension. The remaining pair of suture ends coming through the femur were tied over the femoral cortical button to bring the ACL stump into the scaffold using an arthroscopic surgeon's knot and knot pusher. The arthrotomy was closed in layers and the tourniquet deflated. Sterile dressings, followed by a cold therapy unit (Polar Care, Breg, Carlsbad, CA) and locking hinge knee brace (T-scope, Breg, Carlsbad, CA) were applied. No surgical drain was used.

***Postoperative rehabilitation***

A standardized physical therapy protocol, which did not specify which treatment the patient had received, was provided to all patients. The physical therapists were not informed of the treatment assignment of the patient. For all patients, a locking hinged brace (TScope; Breg, Carlsbad, CA) was applied to limit joint range of motion between 0 to 50 degrees of knee flexion for the first 2 weeks post-operatively, and from 0 to 90 degrees for the next four weeks unless they had a concomitant meniscal repair, in which case the brace range was restricted to 0 to 40 degrees for the first 4 weeks post-operatively before opening the brace up to 0 to 90 degrees of flexion. All patients were provided with a cold therapy unit (Iceman, DJO Global, Vista CA) for post-operative use. Both groups followed the same standardized physical therapy protocol including partial weight bearing for 2 weeks, then weight bearing as tolerated with crutches until 4 weeks post-operatively. Use of a functional ACL brace (CTi brace; OSSUR, Orange County, CA) was recommended from 6 to 12 weeks post-operatively and then for cutting and pivoting sports for 2 years after surgery. Other than the brace use and initial restricted weight bearing, the patients in both groups followed an identical rehabilitation protocol, adapted from that of the Multicenter Orthopaedics Outcomes Network (MOON) ^72; 73^. Phase 1 of the protocol emphasized reducing swelling and regaining full extension (weeks 0 to 2). Phase 2 (~ 2 to 6 weeks) aimed at regaining quadriceps function and a normal gait pattern. Phase 3 (~ 6 to 12 weeks) aimed at progressing neuromuscular function, and Phase 4 (~12 to 18 weeks) focused on running and hopping. Running patterns and jumping began in Phase 5 (~18 to 22 weeks), and patients were gradually progressed to sport specific skill training in Phase 6 (~ 22 to 26 weeks). Patients were cleared for return to sport at the operating surgeon’s discretion after completing an IKDC Subjective Score, hamstring and quadriceps strength measurement and bilateral hop testing at the 6-month visit.

***Non-Imaging predictors***

The following outcome measures were measured following standard clinical protocols:

*International Knee Documentation Committee (IKDC) Subjective Score:* The IKDC Subjective Score is commonly used to assess patient-reported outcomes after ACL surgery ^74-79^. The form used for IKDC scoring assesses the patient’s perception of knee function, symptoms, and sports performance.

*Instrumented AP Knee Laxity Assessment (Knee Arthrometry):* A compuKT 2000 knee arthrometer (MEDmetric Corp, San Diego, CA) was used to measure total translational motion between the tibia relative to the secured femur ^80^. Testing was done according to the MOON protocol in compliance with the manufacturer’s operation manual. Each leg placed on the adjustable thigh support with knee stabilized at 20° to 35° of flexion. The arthrometer was secured to the shank such that the patellar sensor pad was resting on the patella with the knee joint line reference mark on the arthrometer aligned with the subject’s joint line. The ankle and foot were stabilized to limit leg rotation. The examiner applied a posterior (-134N) and anterior (+134N) pressure on an axis perpendicular to the tibia. Displacement (mm) was then documented ^43^. Each test was performed three times and the mean of the three tests was used to obtain a side-to-side difference between the treated and contralateral limbs. An independent examiner performed the tests and knee sleeves were used to cover both knees. The examiner was blinded to the surgical side and study group assignment when performing the physical examination.

*Muscle Strength Assessment:* Hamstring and quadriceps muscle isometric muscle strengths were measured using a hand-held dynamometer (Microfet 2; Hoggan Scientific, LLC, Salt Lake City, UT) that has specifically been validated as a reliable hand-held dynamometer (HHD) in multiple studies ^81-84^. The hamstring strength was measured with the patient prone and the knee in 90° of flexion. The dynamometer was placed on the posterior surface of the lower leg proximal to the ankle. The quadriceps strength was measured with the knee at 90 degrees of flexion with the dynamometer positioned at the distal tibia. An independent examiner performed the tests and knee sleeves were used to cover both knees. The examiner was blinded to the surgical side and study group assignment when performing the physical examination.

*Knee Range of Motion:* Knee passive and active range of motion were measured at each postoperative visit using a goniometer. Knee sleeves were used to cover both knees for all patients, and the examiner was blinded to the surgical side and study group when performing the physical examination. Measurements were done on both knees.

*Functional Hop Test:* Subjects performed single hop aiming for maximum hop distance, and the distance was measured ^85-90^. All tests were performed bilaterally. All measurements were performed in duplicate on each side and the duplicate measurements averaged for further analysis. An independent examiner performed the tests and knee sleeves were used to cover both knees. The examiner was blinded to the surgical side and study group assignment when performing the physical examination.

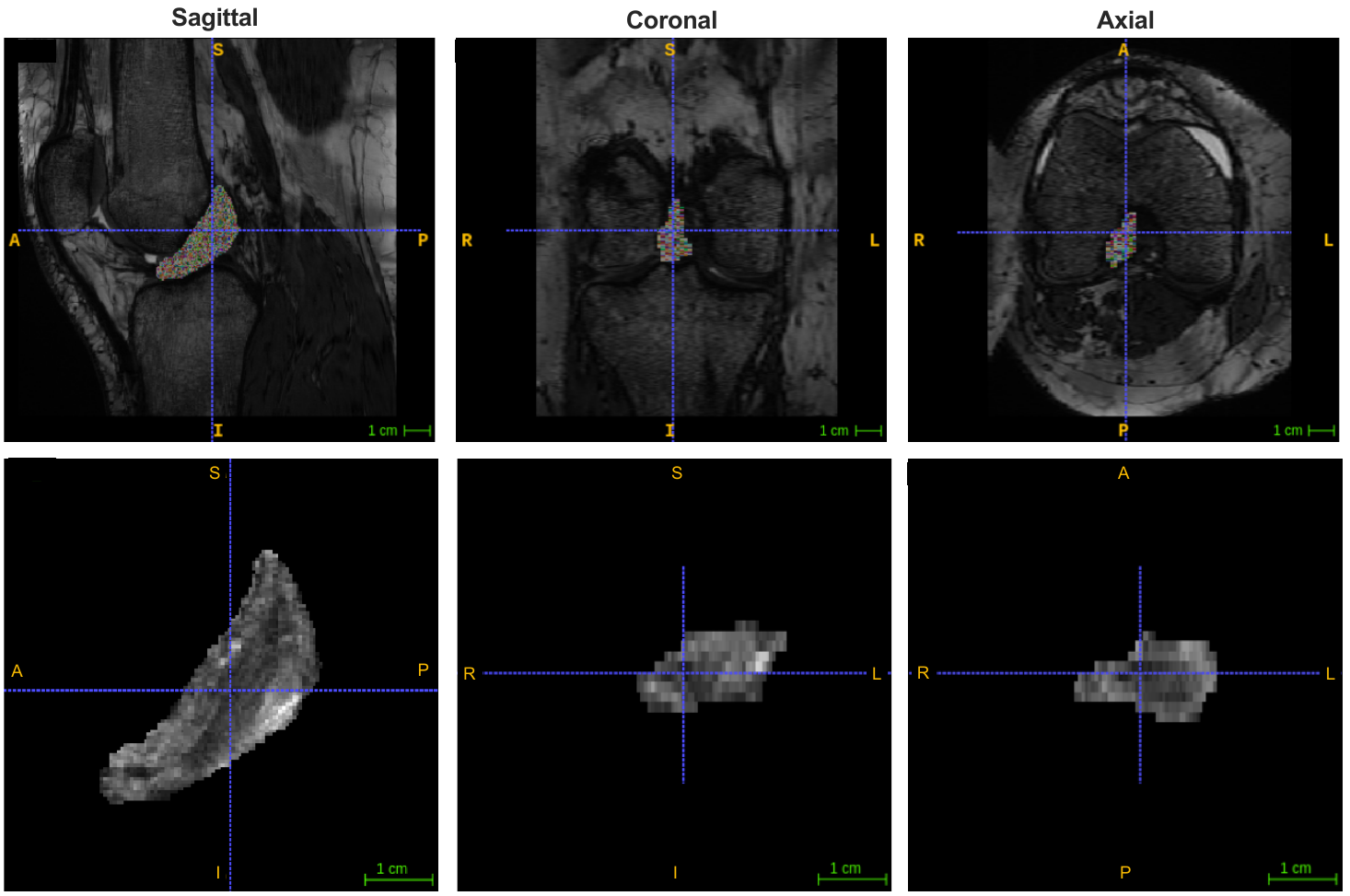

**Figure S1. Multi-planar view of the knee MRI and segmented ACL.** Example of resampled MRI in sagittal, coronal and axial views. The light shaded area indicates ACL.

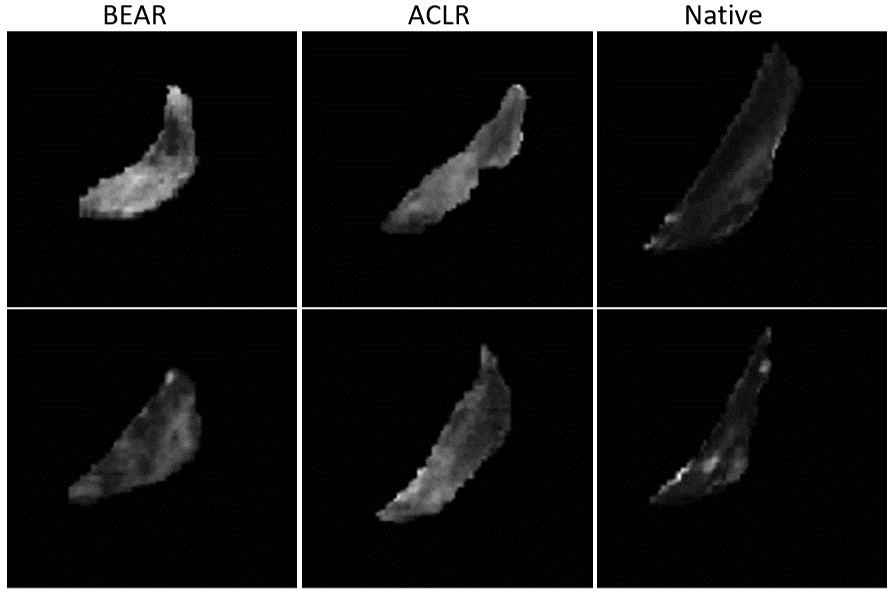

**Figure S2. Segmented ACLs from patients treated with BEAR, ACLR, and intact native ACLs.** Visual similarities between the ACL treated with BEAR and ACLR make ligament type classification a challenging task.

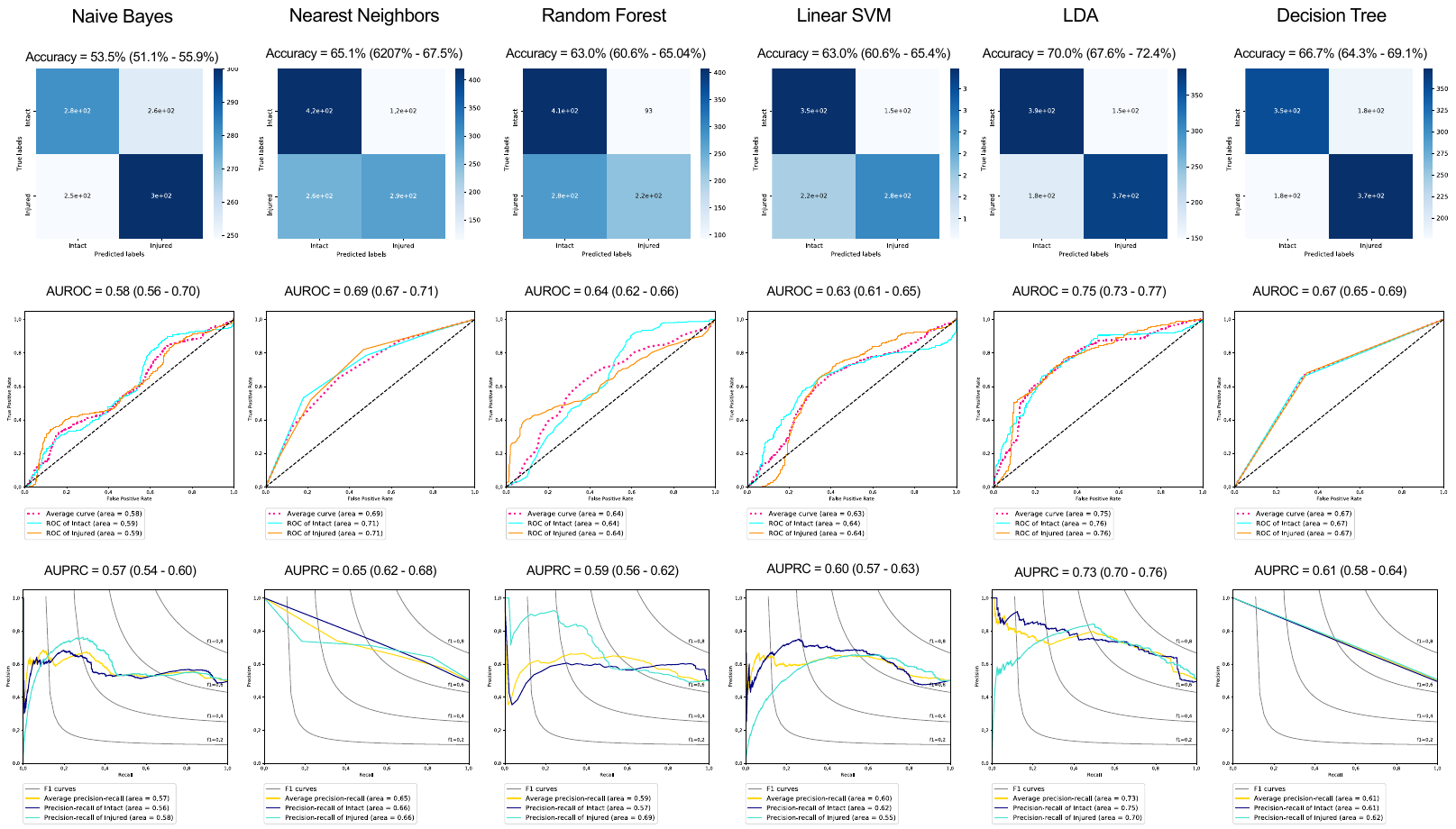

**Figure S3. The model performance metrics for alternative classifiers to distinguish between intact and injured ACLs bases on non-imaging data.** Top row presents the confusion matrices and their corresponding accuracy (95%CI). Middle row presents the receiver operating characteristic curves and their corresponding area under the curve (AUROC) (95%CI). Bottom row presents the precision-recall curves and their corresponding area under the curve (AUPRC) (95%CI).

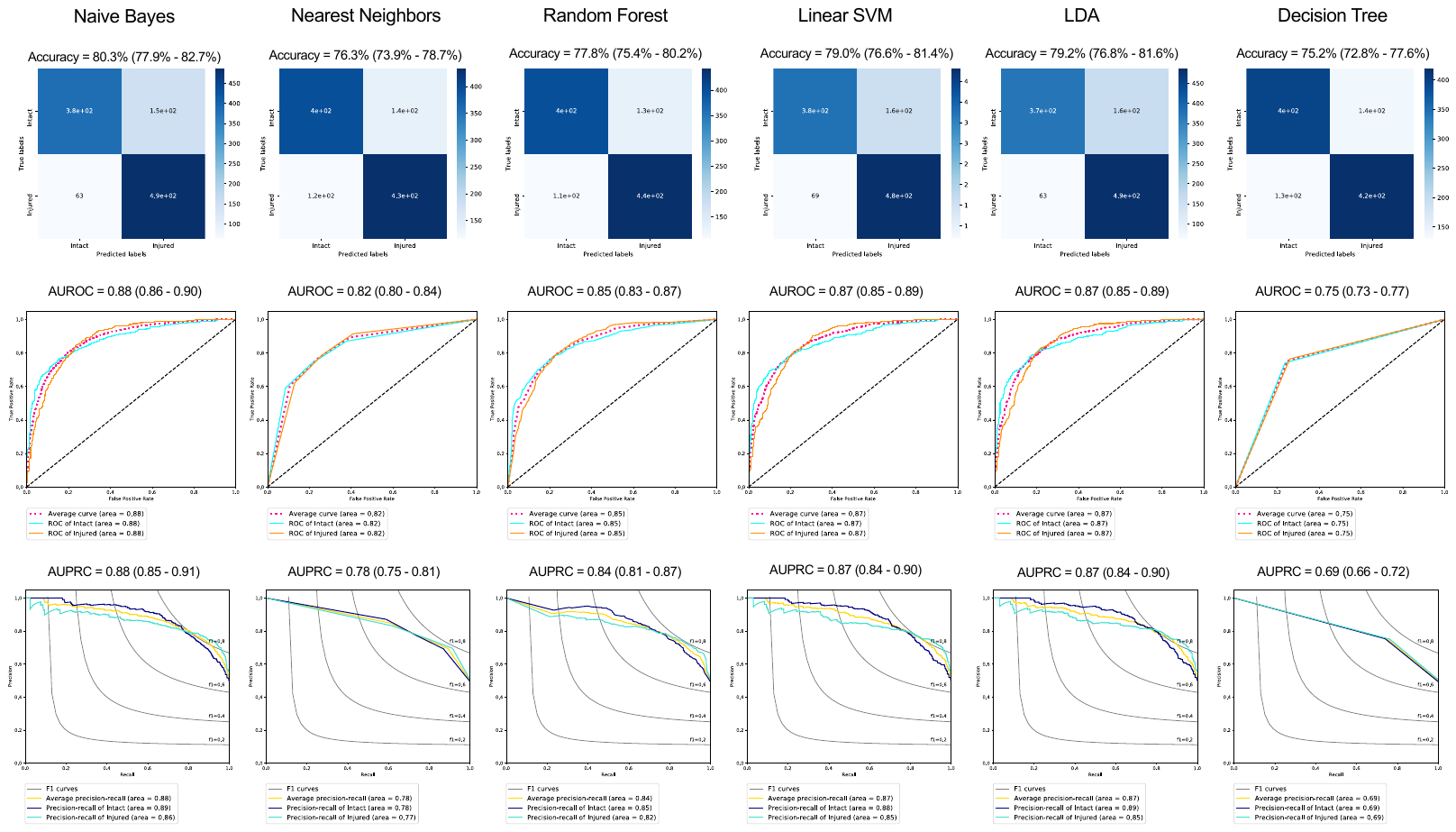

**Figure S4. The model performance metrics for alternative classifiers of multi-modal fusion to distinguish between intact and injured ACLs based on the combination of imaging (MRI segmented ACL and whole-knee MRI) and non-imaging predictors.** Top row presents the confusion matrices and their corresponding accuracy (95%CI). Middle row presents the receiver operating characteristic curves and their corresponding area under the curve (95%CI). Bottom row presents the precision-recall curves and their corresponding area under the curve (95%CI).

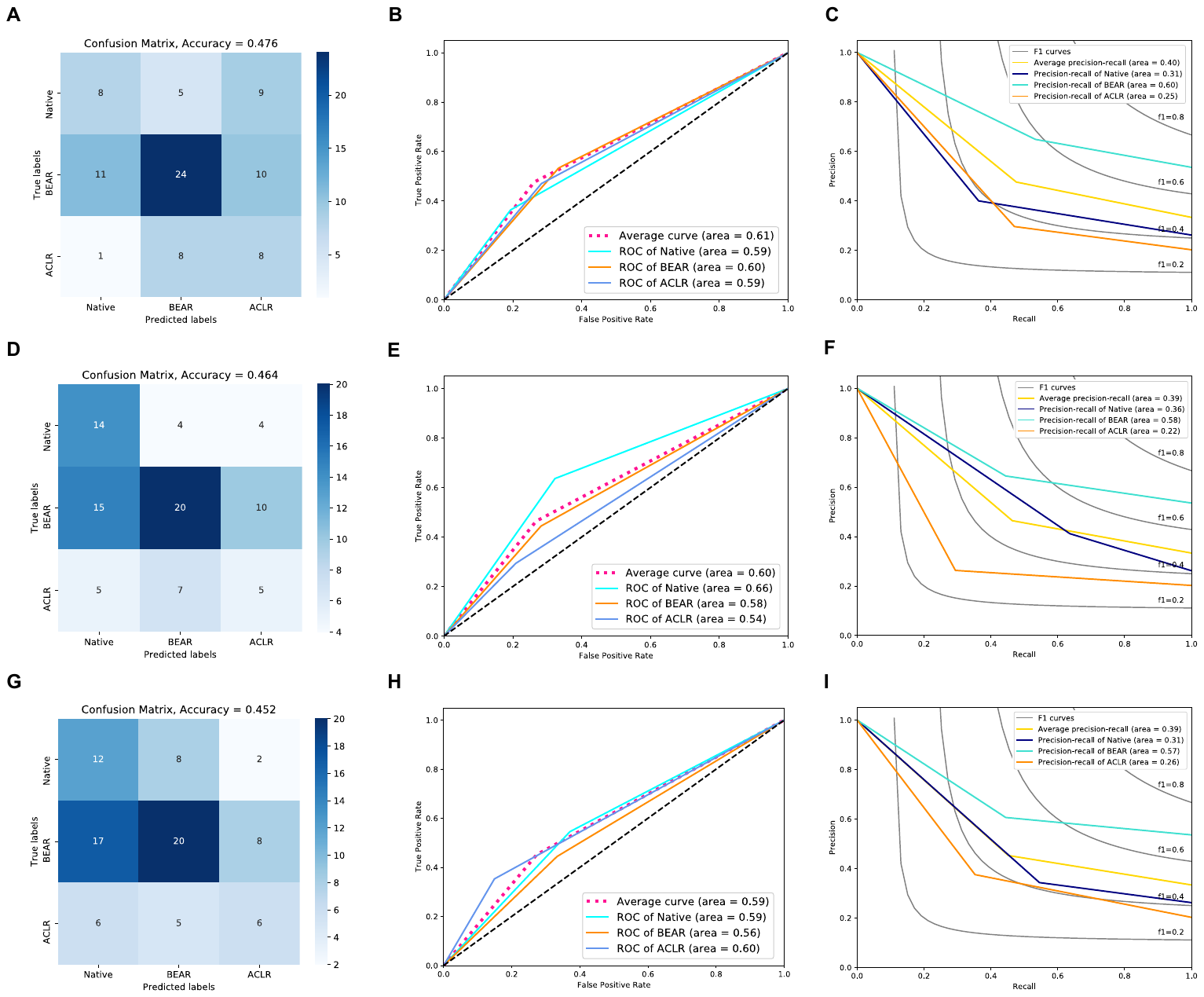

**Figure S5. Human performance in classifying ACL type from segmented ACL.** Each row represents performance metrics from an independent human examiner. **(A, D, and G)** Confusion matrices. **(B, E and H)** Receiver operating characteristic (ROC) curves. **(C, F and I)** Precision recall curves. BEAR: bridge-enhanced ACL restoration, ACL: ACL reconstruction.
